# Relation between motor competence and academic achievement: the mediating role of fitness and cognition in boys and girls

**DOI:** 10.1101/2024.11.21.24317694

**Authors:** Antonio Fernández-Sánchez, Abel Ruiz-Hermosa, Andrés Redondo-Tébar, Ana Díez-Fernández, Vicente Martínez-Vizcaíno, María Eugenia Visier-Alfonso, Mairena Sánchez-López

**Author notes:** E-mail addresses (A. Fernández-Sánchez), (M. Sánchez-López), (A. Ruiz-Hermosa), (A. Redondo-Tébar), (A. Díez-Fernández), (M.E. Visier-Alfonso), (V. Martínez-Vizcaíno). Corresponding author*: Abel Ruiz-Hermosa PhD. ***Authors’ contributions*** Vicente Martínez-Vizcaíno was the principal investigator and obtained the funding. Antonio Fernández-Sánchez was the person who wrote the first draft of the manuscript with the support of Mairena Sánchez-López and Abel Ruiz-Hermosa. Andrés Redondo-Tébar, Ana Díez-Fernández, Vicente Martínez-Vizcaíno and María Eugenia Visier-Alfonso gave statistical and epidemiological support. All authors listed on the manuscript have revised and approved the submission of this version of the manuscript and take full responsibility for the manuscript.

## Abstract

**Introduction:** Good motor competence is linked to better academic achievement in schoolchildren, potentially mediated by fitness and cognition. However, the relative impact of these factors and whether they differ by sex remains unclear. The aim of the present study was to assess the mediating role of fitness components and executive function on this relationship, with a particular focus on sex differences.

**Methods:** This cross-sectional study included 562 schoolchildren (9-11 years, 293 girls) from Cuenca, Spain. We assessed gross motor competence (MABC-2 battery), fitness components (ALPHA battery), executive function (Toolbox battery), academic achievement (mean of language and mathematics grades), weight, height, and sociodemographic variables. Serial multiple mediation models between gross motor competence to AA through fitness and cognition were conducted using the Hayes’ PROCESS SPSS macro, both for the total sample and by sex.

**Results:** Both fitness and executive function partially mediated the relationship between gross motor competence and AA. In the total sample, the direct effect explained most of the total effect (51.22% to 72.68%), followed by the cognitive path (19.51% to 30.73%), fitness path (10.84% to 19.02%), and multiple path (4.43% to 9.27%). In boys, only the cognitive path mediated the relationship (56.35% to 68.91%), while in girls, the fitness path (for cardiorespiratory fitness and speed/agility, 19.08% and 20.27%), the cognitive path (limited to upper body strength, 14.92%), and the multiple path (for cardiorespiratory fitness, speed/agility, and lower body strength, 4.93%, 8.31% and 9.43%) were partial mediators.

**Conclusions:** Our results suggest that part of the effect found between gross motor competence and academic achievement in children occurs from improvements in fitness and executive function. These findings emphasize the need for motor competence programs that target both fitness (especially cardiorespiratory fitness and speed/agility) and executive function to boost academic achievement in children aged 9-11. Additionally, interventions should consider sex-specific differences.

## INTRODUCTION

Motor competence (MC) can be defined as a person’s ability to execute different motor acts, including coordination of fine and gross motor skills that are necessary to manage everyday tasks [1]. There is a growing body of evidence suggesting a positive association between MC and physical health outcomes, including physical fitness and weight status [2]. In addition, MC has been widely recognized as a potential predictor of academic achievement (AA) [3,4]. However, the underlying mechanisms that drive this relationship have not been sufficiently elucidated.

Cognitive processes have been suggested as potential mediators between MC and AA [5]. This association may be attributed to be shared neural pathways and cognitive demands between motor skills and executive function (EF) [6,7]. This is because the same brain regions are partially responsible for the development of MC and EF in childhood [8]. Despite this, few studies have examined the mediating role of EF in this relationship, suggesting that MC exerts a positive influence on AA indirectly through EF [5,9]. However, the evidence on sex differences remains inconclusive. Since one study reported no significant sex differences [10], another found that EF mediated the relationship exclusively in boys [11].

Given the well-established relationship between MC and physical fitness [12], as well as the documented positive association between fitness, cognitive function, and AA [3], it is reasonable to hypothesize that physical fitness could serve as a mediating variable in the relationship between MC and AA. However, to the best of our knowledge, only one study has directly examined this hypothesis, reporting no significant effect [13]. Moreover, while physical fitness is generally positively associated with AA [14], this relationship varies depending on the specific fitness component. Cardiorespiratory fitness (CRF) and speed/agility (S/A) have consistently proven to have a positive effect on AA [15–17], but the role of muscular strength remains unclear [14,15]. Thus, it is reasonable to posit that the mediating effect of fitness on the MC-AA relationship may differ across fitness components.

It thus appears possible to elucidate the association between MC and AA through two principal paths: the cognitive and the fitness path. Furthermore, a third path may be proposed, termed the ‘multiple path’, which suggests that improvements in MC may result in enhancements in physical fitness, which could, in turn, enhance cognitive function and AA. This multiple path will be analyzed for the first time in this study, thereby providing a new and original viewpoint on the relationship between MC and AA.

Finally, research indicates that boys and girls exhibit distinct trajectories in physical and cognitive development [18], as well as in brain activation during cognitive tasks [19]. Such differences could affect the relationship between MC and its connection to fitness, cognitive functions, and AA [20]. Despite these distinctions, there are no studies specifically analyzing the mediating role of fitness and EF by sex.

It is therefore necessary to conduct further in-depth research into the relationships between MC, fitness components, cognitive abilities, and AA, with a particular focus on potential sex differences. The objective of this study was to examine whether fitness and EF act as mediators in the relationship between gross motor competence (GMC) and AA, and to identify potential differences between boys and girls.

## MATERIALS AND METHODS

### Study design and participants

The present study is a cross-sectional analysis of the baseline data from a cluster-randomized controlled trial (NCT03236337) to assess the effectiveness of a physical activity intervention program (MOVI-daFit!) on reducing fat mass and cardiovascular risk and improving physical fitness, EF, and AA among schoolchildren. Ten schools from ten towns in the province of Cuenca, Spain, participated in the study. In all schools, all children belonging to the fourth and fifth grades of primary school (9–11 years old) were invited to participate. The study design, sampling procedures and methods have been fully described elsewhere [21].

The study protocol was approved by The Clinical Research Ethics Committee of the ‘Virgen de la Luz’ Hospital in Cuenca (REG: 2016/PI021). The approval of the principal and the board of governors was sought from each school. The children were informed about the study’s objectives and gave their verbal consent to participate, with the principal, researchers, and their peers as witnesses. Additionally, the parents or legal guardians of all participants provided written informed consent.

### Instruments and study variables

The recruitment period for this study began on September 11, 2017, and ended on October 26, 2017. The measurements were conducted by researchers who were trained in advance to standardize the procedures and were blinded to the participants’ group assignments.

### Gross motor competence

Gross motor competence refers to the degree of skilled performance in a wide range of motor tasks as well as the movement coordination and control underlying a particular motor outcome which involves the larger muscle groups [22] and is a prerequisite for child physical activity participation and also for engagement in learning and social activities, including sports and games [23]. To assess GMC, two dimensions (aiming-catching and balance skills) of the validated Spanish version of the Movement Assessment Battery for Children-Second Edition (MABC-2) were used [1] for the age ranges 2 (7-10 years) and 3 (11-16 years). The tasks performed were: 1) Aiming-catching 1 *(catching)*: The child had to throw a tennis ball against the wall and catch it with both hands (9-10 years); The child had to throw a tennis ball against the wall and then catch it using only one hand (11 years). 2) Aiming-catching 2 *(throwing)*: The child had to aim a beanbag into a red circle on a mat (9-10 years); The child had to throw a tennis ball into a red circle on the wall (11 years). 3) Balance 1 *(static)*: Single-board balance - The child had to balance on one foot on the balance board (9-10 years); Two-board balance - The child had to balance on the balance board, ensuring that the heel of one foot and the toes of the other foot touch (11 years). 4) Balance 2 *(dynamic)*: Heel to toe - The child had to walk along a line while the heel of one foot touches the toes of the other foot. (9-10 years); Walking backwards heel to toe - The child had to walk backwards along a line, making sure that the toes of one foot touch the heel of the other (11 years). 5) Balance 3 *(dynamic)*: Hopscotch - The child had to jump forward on one leg from mat to mat starting from a standing position (9-10 years); Zigzag hopping - The child had to jump diagonally from one mat to another on one leg (11 years) [24]. Each test received a raw score (number of catches, number of hits on target, seconds on one leg, number of steps on the line and number of right jumps, respectively) where a higher score indicated a better GMC. The resulting variable, *GMC Total Score,* was calculated as the sum of the z-scores for aiming-catching, static balance, and dynamic balance.

### Fitness variables

Fitness is defined as a set of attributes related to a person’s ability to perform physical activities that require aerobic capacity, endurance, strength, or flexibility and is determined mostly by a combination of regular activity and genetically inherited ability [25]. The evidence-based ALPHA-Fitness test battery [26] was used, including the following tests:

*Cardiorespiratory fitness,* using the 20-metre shuttle run test, a valid test to estimate aerobic capacity in children. The children were required to run 20 meters between two lines while keeping pace with the rhythm marked by acoustic signals. The initial speed of 8.5 km/h was increased by 0.5 km/h for each stage (one stage equals one minute). The last half stage completed by the child was considered an indicator of CRF. Maximal oxygen intake (VO_2_max) was estimated by applying the Léger formula [27].

*Speed-agility*, using the 4×10 meter shuttle run test. Children were required to run and turn (shuttle) 10 meters twice at maximum speed (4×10m). Two parallel lines were drawn on the floor. When the start was given, the children ran as fast as possible to the other line and returned to the starting line twice, crossing both lines with both feet. Two attempts were made, with an interval of five minutes between attempts, and the best score was recorded in seconds. A lower score indicated a better S/A.

*Upper Body strength* (UBS), through the handgrip strength test, determined in kilograms using a digital dynamometer with adjustable grip TKK 5401 Grip-DW (Takeya, Tokyo, Japan). The handgrip strength test has high-to-very high construct validity with total muscle strength in healthy children, among other parameters [28]. The test was performed twice with each hand and the arithmetic mean of the four measurements was computed. Handgrip data was normalized by dividing absolute handgrip strength (kg) by body weight (kg).

*Lower Body strength* (LBS), using the standing broad jump test, which measures explosive LBS. The children jumped horizontally to achieve the maximum distance and the best of three attempts was recorded in centimeters.

### Executive functions

Executive functions refer to a family of mental processes that have been categorized in three core functions: inhibitory control (the ability to selectively attend, focusing on what we choose and suppressing attention to other stimuli), cognitive flexibility (the ability to change perspectives) and working memory (the ability to hold information in mind and mentally working with it) [29]. Three EFs, using the NIH Toolbox software (NIH Toolbox in Spanish, v. 1.8) [30] were assessed. All tests were performed individually in a quiet classroom using a tablet (iPad Pro, Apple, Inc., California, USA). The evaluation tests included in the NIH Toolbox for each cognitive domain are listed as follows:

*Inhibitory control,* using an adaptation of the Flanker test [31]. Participants had to indicate the left-right direction of a centrally shown stimulus while inhibiting their attention to the potentially incongruent stimuli around it. In some trials, the orientation of the flanking stimuli was congruent with the orientation of the central stimulus (> > > > > or < < < < <,) while in other trials, this orientation was incongruent (> > < > > or < < > < <). The NIH Toolbox contained a practice block of trials. If participants passed it, a 20-trial block was presented. These trials consisted of a succession of congruent and incongruent combination of arrows. Using a two-vector method that included both reaction time and accuracy, a final score was calculated for school children who reached a high level of accuracy (>80%), which was as follows: (0.25 x number of correct responses) + 5 – log_10_ [(congruent reaction time + incongruent reaction time) / 2]. A total score considering accuracy was determined for children who scored less than 80%.

*Cognitive flexibility*, using the Dimension Change Card Sort (DCCS) [31]. Participants were shown two target cards and asked to order a group of bivalent test cards, first according to one dimension and then according to the other dimension. After a four-trial practice, children were given a 30-trial block with both ‘shape’ and ‘color’ requirements. Using reaction time and accuracy percentage on preswitch and postswitch, a raw score was determined. Using a two-vector method that integrated both accuracy and reaction time, a final score was calculated for children who reached a high level of accuracy (>80%) as follows: (0.167 x number of correct responses) + 5 – log_10_ [(congruent reaction time + incongruent reaction time) / 2]. For participants scoring <80%, a total outcome considering accuracy was determined.

*Working memory*, using the List Sorting Working Memory Test [32]. Children were given a group of pictures, and items were presented both auditorily and visually. Then, they were required to repeat the names of the items observed in order of size, from smallest to biggest. The number of items shown in each trial increased by one for each series. This test was made up of two parts. In the first one, all the items belonged to the same category (animals or food items). In the second one, the items from both categories (food items and animals) were presented together, and children had to repeat the items by category and size. The List Sorting “Total Score” was composed of final scores based upon a sum of the total correct trials across the two lists.

The resulting variable, *EF Total Score*, was calculated as the sum of the z-scores for inhibitory control, cognitive flexibility, and working memory.

### Academic achievement

The final grades in language and mathematics, provided by the school administration, were used to determine AA. These subjects were selected because, in Spain, they are considered instrumental subjects, as they serve as a foundation for learning other subjects and help in everyday life and activities. The children’s final grades from the previous academic year were used (range from 1 to 10 score, with 10 being the best grade). The arithmetic mean of the grades in language and mathematics was used in the analysis. In Spain, final grades represent the student’s work across a complete academic year.

### Anthropometry

Weight was measured using a scale (Seca 861, Vogel and Halke, Hamburg, Germany) with the child in light clothing and barefoot. For height, a wall stadiometer (Seca 222, Vogel and Halke) was used with children barefoot and standing upright with their sagittal midline in contact with the backboard. Both weight and height were measured twice, and their arithmetic mean was considered for the analysis. Body mass index (BMI) was calculated as weight (kg) / height (m^2^).

### Sociodemographic data

The age, sex and mother’s education level were gathered using a questionnaire administered to the parents. Data on mother’s education level as the maximum level of education attained by the children’s mothers gathered the validated scale proposed by the Spanish Society of Epidemiology [33] to measure socioeconomic status. This questionnaire includes an item referred to mother’s education with six response options: i) no literacy skills; ii) no studies; iii) elementary studies; iv) secondary studies; v) high school; and vi) university studies. In our study, these six categories were collapsed into three, due to the small number of individuals in the lower and upper categories: lower/lower middle (no literacy skills, no studies, and elementary studies), middle (secondary studies) and upper middle/upper (high school and university studies). The mother’s education level was used as covariate in the mediation models (as an interval variable), because it has been shown to be a strong predictor of children’s AA [34].

### Statistical analysis

Interval variables were summarized as means and standard deviations. The mother’s education level was expressed as counts and frequencies. Both statistical (Kolmogorov-Smirnov test) and graphical procedures (normal probability plots) were used to evaluate the goodness of fit of variables to a normal distribution and all variables fitted to a normal distribution. Differences in all variables between boys and girls were tested using Student’s t-tests.

Pearson correlation coefficients were estimated to examine the relationship between GMC, CRF, S/A, UBS, LBS, EF Total Score, and AA for the total sample and by sex. Values below 0.1 were defined as trivial, weak in the range 0.1–0.29, moderate in the interval 0.3–0.49, and large when greater than 0.5 [35].

The mediation effect of different fitness components (CRF, S/A, UBS, and LBS) and EF on the relationship between GMC and AA for the total sample and by sex was tested using the PROCESS SPSS Macro, version 4.2, selecting model 6 (serial multiple mediation analysis) and 10.000 bias-corrected bootstrap samples [36]. For this analysis, GMC was entered in the model as the independent variable, the proposed mediators were, consecutively, fitness variables and EF, and AA was included as the dependent variable. Each fitness component was individually entered in a different model: model a, CRF; model b, S/A; model c, UBS; and model d, LBS. In the mediation model, the total (c), and direct effects (a_1_, a_2_, b_1_, b_2_, d_12_ and ć) were estimated. Additionally, this model examines three indirect effects (IEs) (fitness path, cognitive path, and multiple path) that indicate the change in AA for each unit change in GMC that is mediated by each fitness variable and EF. The IEs were considered significant when the 95% confidence interval did not include zero.

The IE is represented by the path from GMC to AA via mediators (paths a, b, and d). The fitness path (a_1_-b_1_), from GMC to AA via fitness variables (a, CRF; b, S/A; c, UBS; and d, LBS). The cognitive path (a_2_-b_2_), from GMC to AA via EF. Finally, the multiple path (d_12_), from GMC to AA via fitness variables (a, CRF; b: S/A; c, UBS; and d, LBS) and EF. Proposed cut points to quantify effect size were 0.14, 0.36, and 0.51 for small, medium and large effect sizes, respectively [37]. The percentages of mediation (P_M_) were calculated as (IE/total effect) x 100 to estimate the percentage of the total effect explained by the mediation paths. All analyses were adjusted by age, BMI, and motheŕs education level.

Data analyses were conducted using IBM SPSS Statistics version 28.0 (IBM® SPSS®, Armonk, NY: IBM Corp.), and the level of significance was set at p < 0.05.

## RESULTS

The characteristics of the total sample, as well as by sex, are presented in **Table 1**. Of the 562 schoolchildren involved in the study, 293 were girls (52.14%), with a mean age of 10.02 years (SD = ± 0.71). Boys scored significantly higher on aiming-catching (p < 0.001), CRF (p < 0.001), S/A (p = 0.006), UBS (p = 0.004), and LBS (p < 0.001) while girls scored significantly higher on static and dynamic balance (p < 0.001), and cognitive flexibility (p = 0.014).

**Table 1.**
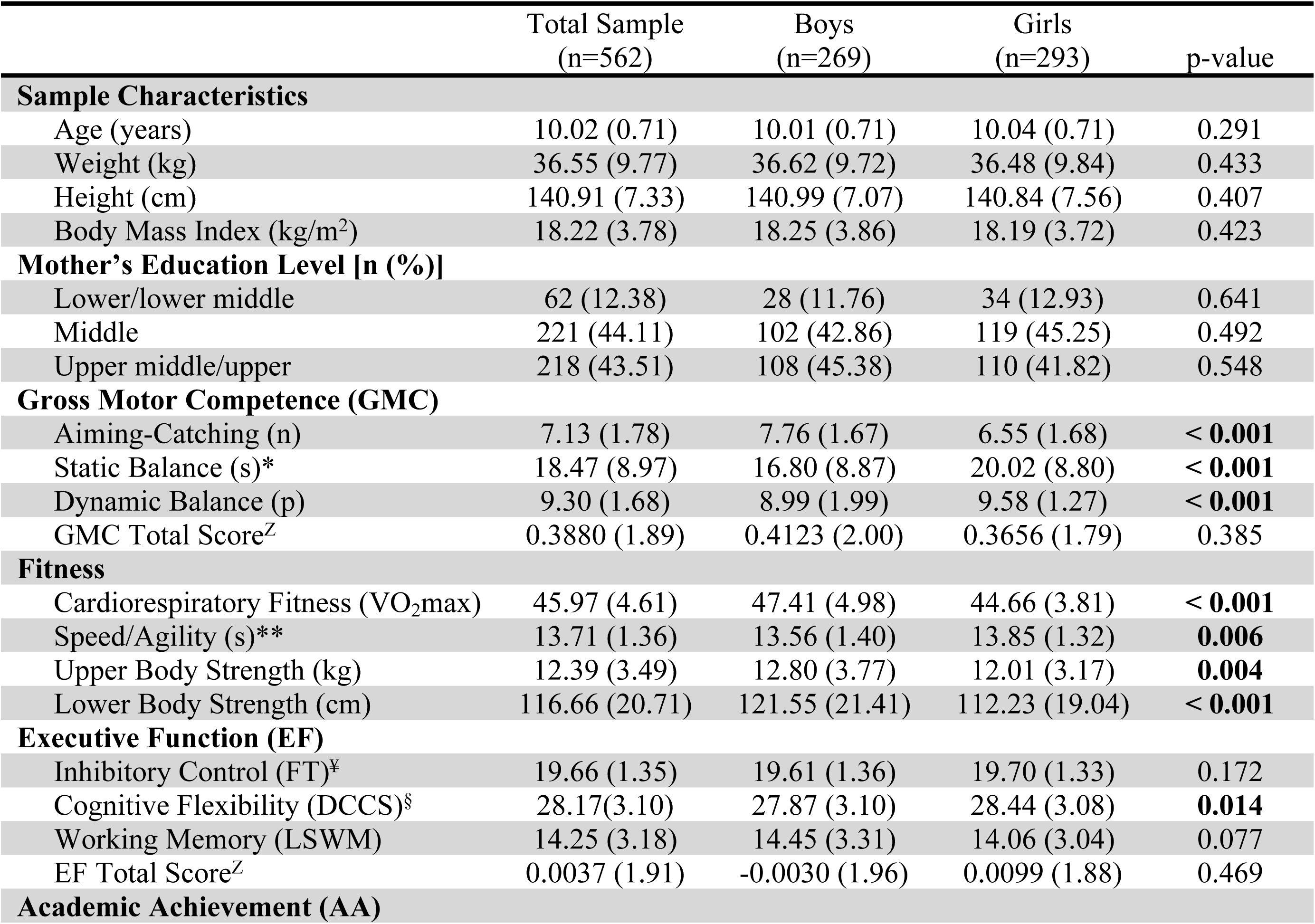
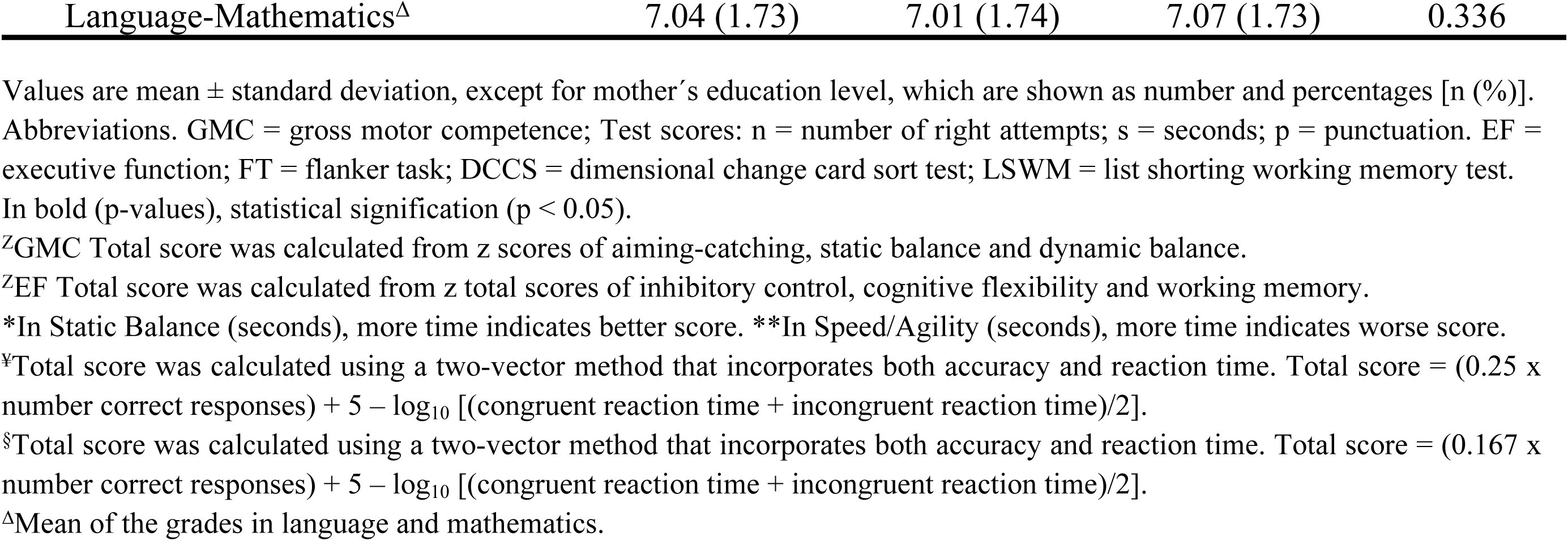
Characteristics of the study sample by sex.

|  | Total Sample<br>(n=562) | Boys<br>(n=269) | Girls<br>(n=293) | p-value |
| --- | --- | --- | --- | --- |
| **Sample Characteristics** |  |  |  |  |
| Age (years) | 10.02 (0.71) | 10.01 (0.71) | 10.04 (0.71) | 0.291 |
| Weight (kg) | 36.55 (9.77) | 36.62 (9.72) | 36.48 (9.84) | 0.433 |
| Height (cm) | 140.91 (7.33) | 140.99 (7.07) | 140.84 (7.56) | 0.407 |
| Body Mass Index (kg/m<sup>2</sup>) | 18.22 (3.78) | 18.25 (3.86) | 18.19 (3.72) | 0.423 |
| **Mother's Education Level [n (%)]** |  |  |  |  |
| Lower/lower middle | 62 (12.38) | 28 (11.76) | 34 (12.93) | 0.641 |
| Middle | 221 (44.11) | 102 (42.86) | 119 (45.25) | 0.492 |
| Upper middle/upper | 218 (43.51) | 108 (45.38) | 110 (41.82) | 0.548 |
| **Gross Motor Competence (GMC)** |  |  |  |  |
| Aiming-Catching (n) | 7.13 (1.78) | 7.76 (1.67) | 6.55 (1.68) | < **0.001** |
| Static Balance (s)* | 18.47 (8.97) | 16.80 (8.87) | 20.02 (8.80) | < **0.001** |
| Dynamic Balance (p) | 9.30 (1.68) | 8.99 (1.99) | 9.58 (1.27) | < **0.001** |
| GMC Total Score<sup>Z</sup> | 0.3880 (1.89) | 0.4123 (2.00) | 0.3656 (1.79) | 0.385 |
| **Fitness** |  |  |  |  |
| Cardiorespiratory Fitness (VO<sub>2</sub>max) | 45.97 (4.61) | 47.41 (4.98) | 44.66 (3.81) | < **0.001** |
| Speed/Agility (s)** | 13.71 (1.36) | 13.56 (1.40) | 13.85 (1.32) | **0.006** |
| Upper Body Strength (kg) | 12.39 (3.49) | 12.80 (3.77) | 12.01 (3.17) | **0.004** |
| Lower Body Strength (cm) | 116.66 (20.71) | 121.55 (21.41) | 112.23 (19.04) | < **0.001** |
| **Executive Function (EF)** |  |  |  |  |
| Inhibitory Control (FT)<sup>‡</sup> | 19.66 (1.35) | 19.61 (1.36) | 19.70 (1.33) | 0.172 |
| Cognitive Flexibility (DCCS)<sup>§</sup> | 28.17(3.10) | 27.87 (3.10) | 28.44 (3.08) | **0.014** |
| Working Memory (LSWM) | 14.25 (3.18) | 14.45 (3.31) | 14.06 (3.04) | 0.077 |
| EF Total Score<sup>Z</sup> | 0.0037 (1.91) | -0.0030 (1.96) | 0.0099 (1.88) | 0.469 |
| **Academic Achievement (AA)** |  |  |  |  |
| Language-Mathematics<sup>Δ</sup> | 7.04 (1.73) | 7.01 (1.74) | 7.07 (1.73) | 0.336 |

Values are mean ± standard deviation, except for mother's education level, which are shown as number and percentages [n (%)]. Abbreviations. GMC = gross motor competence; Test scores: n = number of right attempts; s = seconds; p = punctuation. EF = executive function; FT = flanker task; DCCS = dimensional change card sort test; LSWM = list shorting working memory test. In bold (p-values), statistical signification (p < 0.05). <sup>Z</sup>GMC Total score was calculated from z scores of aiming-catching, static balance and dynamic balance. <sup>Z</sup>EF Total score was calculated from z total scores of inhibitory control, cognitive flexibility and working memory. <sup>\*</sup>In Static Balance (seconds), more time indicates better score. <sup>\*\*</sup>In Speed/Agility (seconds), more time indicates worse score. <sup>‡</sup>Total score was calculated using a two-vector method that incorporates both accuracy and reaction time. Total score = (0.25 x number correct responses) + 5 - log<sub>10</sub> [(congruent reaction time + incongruent reaction time)/2]. <sup>§</sup>Total score was calculated using a two-vector method that incorporates both accuracy and reaction time. Total score = (0.167 x number correct responses) + 5 - log<sub>10</sub> [(congruent reaction time + incongruent reaction time)/2]. <sup>Δ</sup>Mean of the grades in language and mathematics.

The Pearson’s correlation coefficients for the total sample, as well as by sex, are shown in **Table 2**. For the total sample, all the fitness variables showed a significant correlation with AA and EF (r values between 0.131 and 0.249, p < 0.01), except between UBS and EF. By sex, all the fitness variables show a significant correlation with AA and EF except between UBS and EF for both sexes, between UBS and AA for girls, and between LBS and AA for boys. In all the fitness variables, the correlation coefficient with both EF and AA was consistently higher in girls compared to boys. Specifically, S/A stands out as the fitness variable with the strongest correlation with EF (r = -0.217 in boys and r = -0.269 in girls; p < 0.01), while CRF exhibits the highest correlation with AA (r = 0.219 in boys and r = 0.317 in girls; p < 0.01). Most of the mentioned associations show weak correlation strength. However, notable moderate strength correlations are observed between CRF and AA (r = 0.317; p < 0.01), as well as between LBS and EF (r = 0.312; p < 0.01), both exclusively in girls.

**Table 2.**
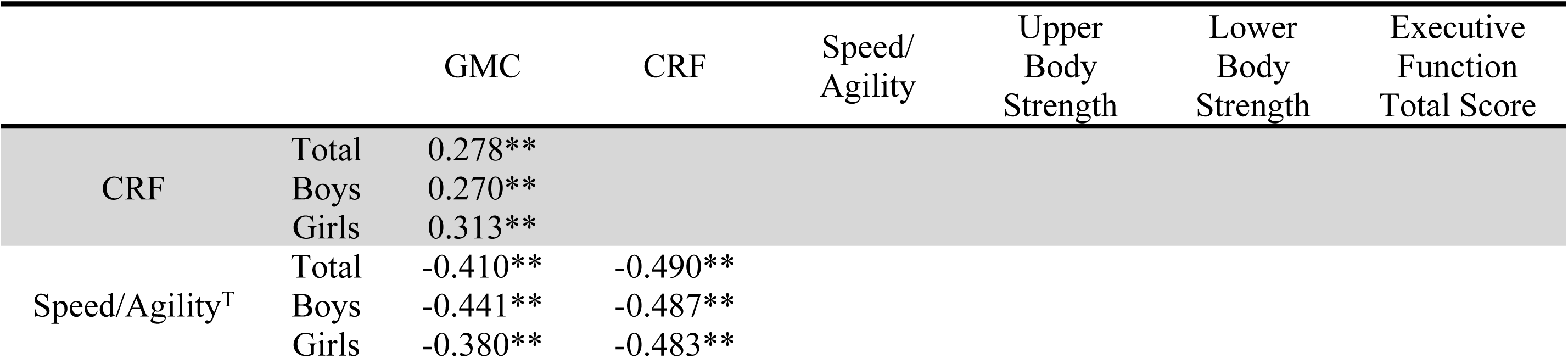
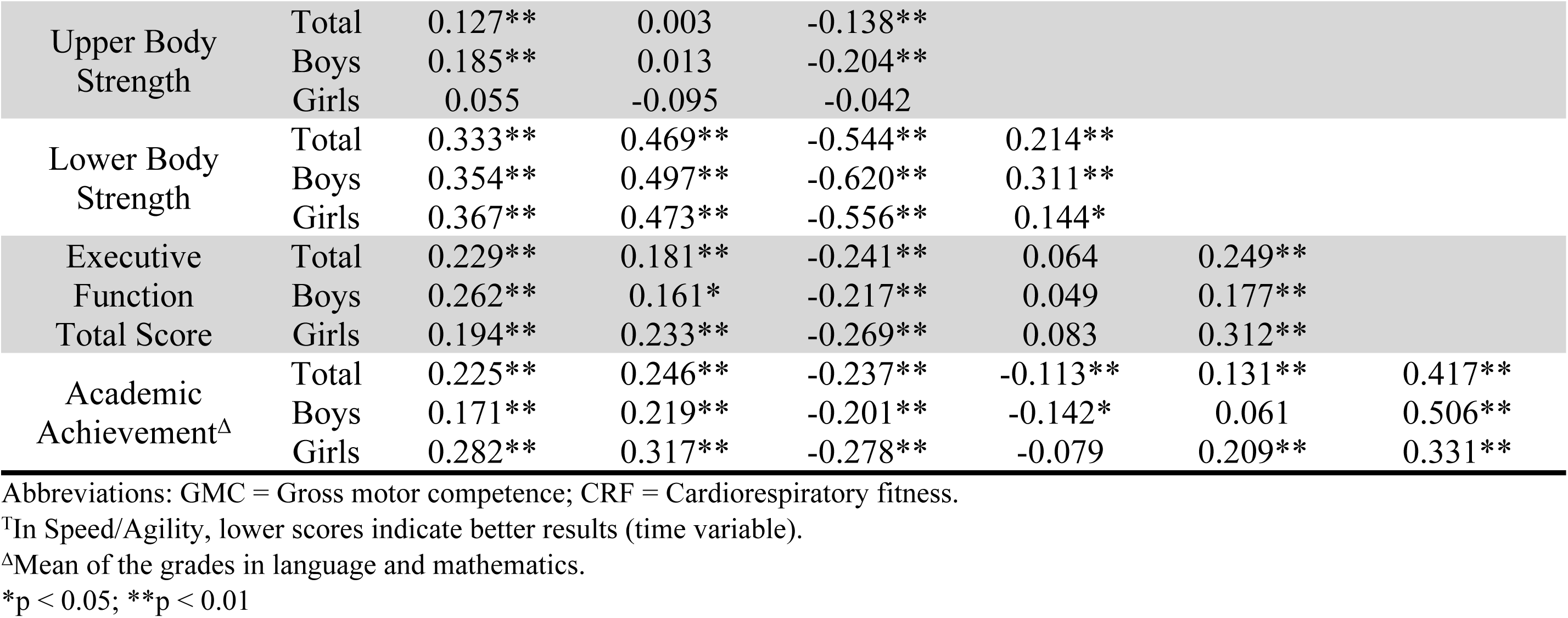
Bivariate correlation coefficients among the study variables for the total sample and by sex.

Multiple mediation models for the total sample indicated a consistent association between GMC and AA across all models, with unstandardized coefficients ranging from 0.203 to 0.205 (p<0.001). Most of this effect was explained by the direct association, with coefficients between 0.105 and 0.149 (p<0.05). Additionally, all three indirect paths (fitness, cognitive, and multiple) were significant for all fitness variables, except for the fitness path in UBS and LBS, and the multiple path in S/A and UBS **(Fig 1)**.

**Fig 1.**
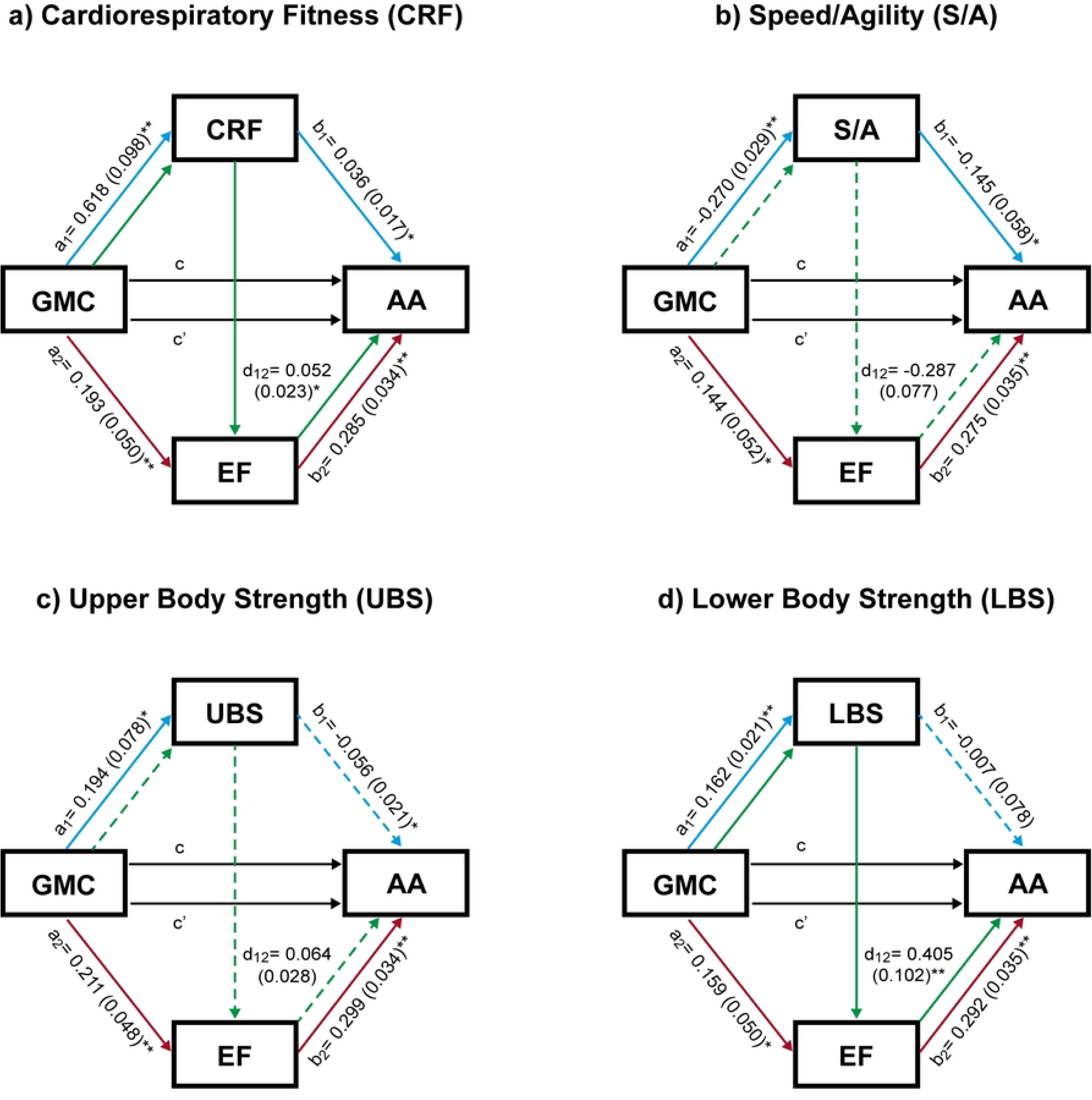
Serial multiple mediation model. (**total sample**). Association between gross motor competence (independent variable) and academicachievement (dependent variable), with different fitness components (CRF, S/A, UBS and LBS) and executive functions as mediators, controlling for age, body mass index and mother’s education level. Values for the a_1_, a_2_, b_1_, b_2_ and d_12_ paths are expressed as the unstandardized regression coefficients (standard error). Values c and c’ express total and direct effect, respectively. GMC gross motor competence, CRF cardiorespiratory fitness, S/A speed/agility, UBS upper body strength, LBS lower body strength, EF executive function, AA academic achievement.

Among boys (**Fig 2)** the total effect of GMC on AA was significantly lower, with coefficients ranging from 0.119 to 0.130 (p<0.05) highlighting no direct association between GMC and AA for any of the variables studied. Instead, improvements in AA through GMC consistently occurred via EF (cognitive path). In contrast, among girls, the total effects were significantly higher (0.295 to 0.304, p<0.001), with most of the effect explained by the direct association between GMC and AA, which was significant across all models, with coefficients ranging from 0.200 to 0.252 (p<0.05). Additionally, for CRF and S/A, both the fitness and multiple paths were significant, while only the cognitive path was significant for UBS, and only the multiple path was significant for LBS.

**Fig 2.**
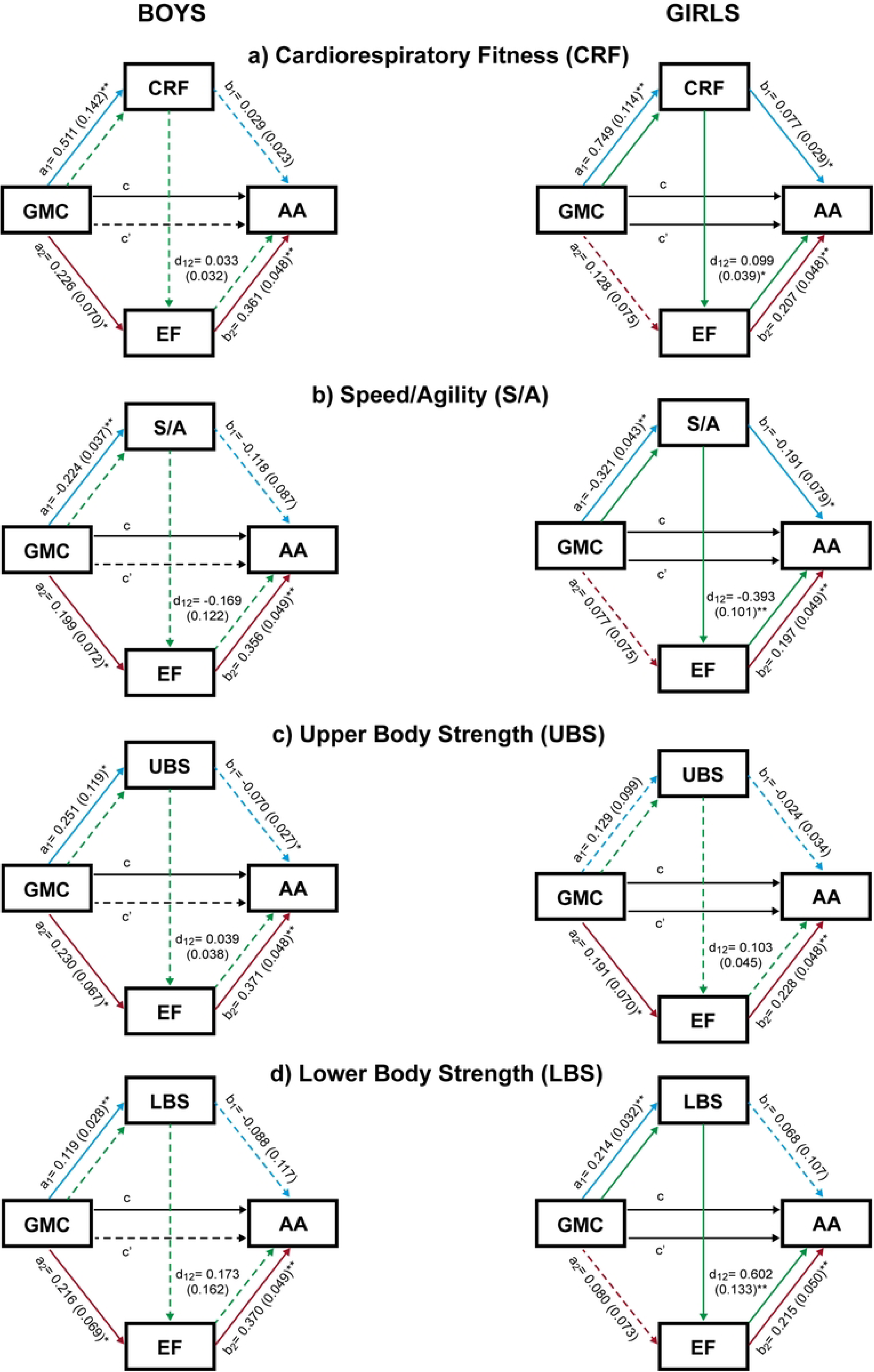
Serial multiple mediation model. (**boys and girls**). Association between gross motor competence (independent variable) and academic achievement (dependent variable), with different fitness components (CRF, S/A, UBS and LBS) and executive functions as mediators, controlling for age, body mass index and mother’s education level. Values for the a_1_, a_2_, b_1_, b_2_ and d_12_ paths are expressed as the unstandardized regression coefficients (standard error). Values c and c’ express total and direct effect, respectively. GMC gross motor competence, CRF cardiorespiratory fitness, S/A speed/agility, UBS upper body strength, LBS lower body strength, EF executive function, AA academic achievement.

The total, direct, and IEs of the different paths from the serial multiple mediation analyses, along with the mediation percentages for direct and IEs, are summarized in **Table 3**. Across all models examined, a positive association between GMC and AA was observed. For the total sample, most of the total effect was accounted for the direct effect (51.22% to 72.68%), followed by the IEs of the cognitive path (19.51% to 30.73%), fitness path (10.84% to 19.02%), and multiple path (4.43% to 9.27%). In boys, more than half of the total effect was explained by the IE of the cognitive path (56.35% to 68.91%). In contrast, in girls, the majority of the total effect was attributed to the direct effect of GMC on AA (66.45% to 85.42%), followed by the IEs of the fitness path (19.08% to 20.27%), cognitive path (for UBS only, 14.92%), and multiple path (4.93% to 9.43%).

**Table 3.**
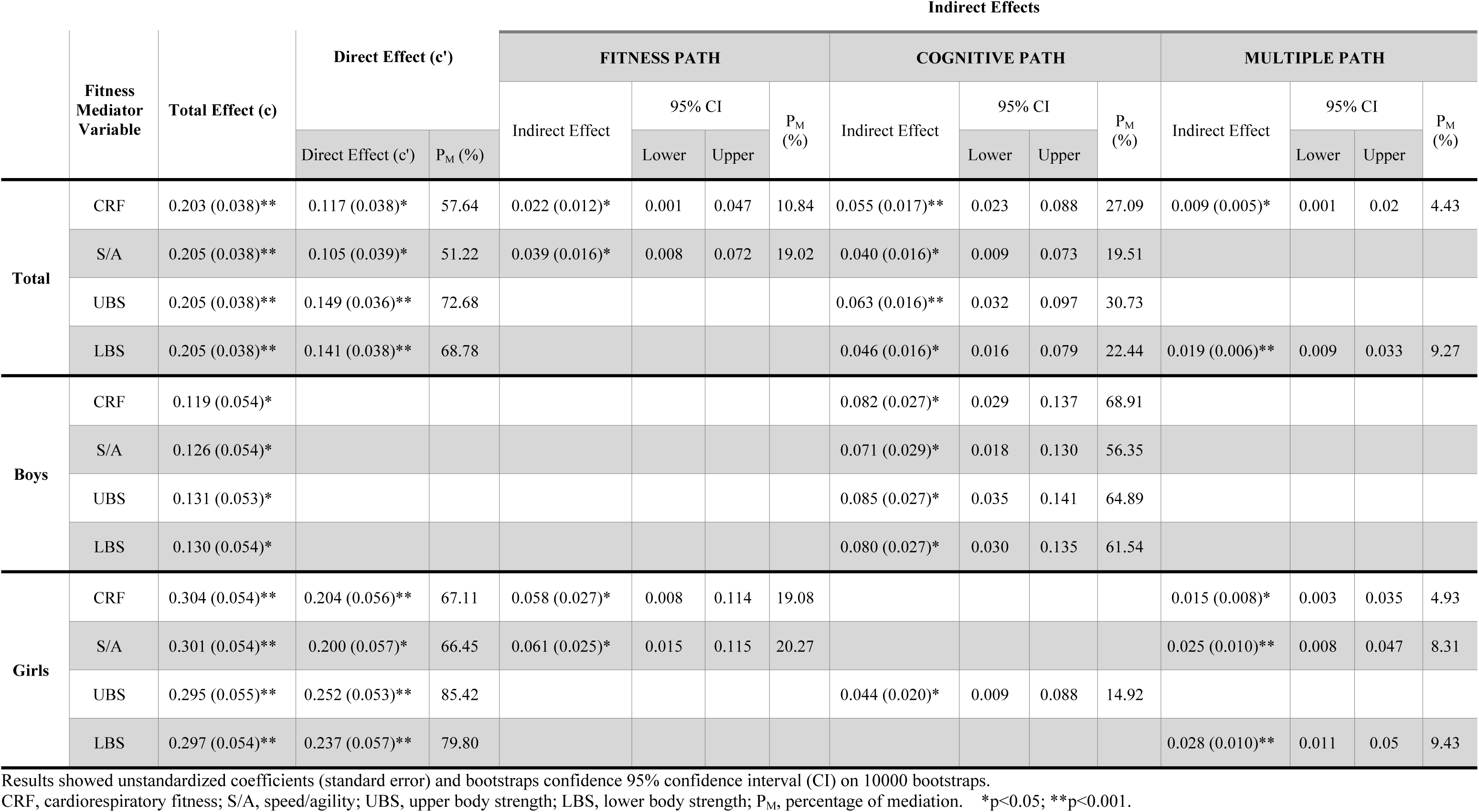
Total, direct, and indirect effects of the different paths from the serial multiple mediation analyses, investigating fitness and executive functions as mediators between gross motor competence and academic achievement, controlling for age, body mass index, and mother’s education level.

## DISCUSSION

To our knowledge, this is the first study to use multiple mediation models to examine the influence of physical fitness components and EF on the relationship between GMC and AA. Our analyses indicate that the fitness path (for CRF and S/A), the cognitive path (across all fitness components), and the multiple path (for CRF and LBS) partially mediated the relationship between GMC and AA in primary schoolchildren even after controlling for age, BMI and mother’s education level. Moreover, significant sex differences were also found, with EF being the only mediator of this relationship in boys, whereas fitness variables, particularly CRF and S/A, were found to be the main mediators in girls.

Compared to previous studies, our findings agree with those obtained by Schmidt et al. [9], who reported that motor ability predicted AA fully mediated through EF performance in a longitudinal study with children aged 10 to 12 years using structural equation models, and with a previous study by our group that found that different domains of EF acted as a partial mediator in this relationship [11]. Similarly, findings by Cadoret et al. [5], support a mediation model in which the relationship between motor proficiency and AA is mediated by cognitive ability in 7-year-old children.

To our knowledge, only one study [13] has investigated the mediating role of CRF between MC and AA in Finnish schoolchildren, which limits the comparability of our results. This study examined whether parent-reported MC at 8 years, predicts AA (grades) via CRF (cycle ergometer test) at 16 years. In contrast to our data, this study found that CRF did not mediate this relationship. These differences could be explained by the different instruments and predictive methodologies used to evaluate CRF, MC, and AA, as well as changes in educational level and the transformations that occur during puberty. However, further studies are needed to gain a deeper understanding of the mediating role of fitness in the MC-AA relationship.

Our findings also suggest that improvements in GMC lead to increased physical fitness, which, in turn, enhances EF, ultimately improving AA in schoolchildren. This multiple mediation path showed that improving AA through fitness and EF was effective for CRF and LBS in the total sample, and for CRF, S/A, and LBS in girls. These findings highlight the importance of designing motor activities/games that improve both the components of physical fitness (especially CRF and S/A) and the mental skills of EF in programs focused on GMC to improve AA in children aged 9-11 years.

Our study found distinct sex differences in the relationship between GMC and AA. For girls, this relationship is complex, with direct links and several mediating paths through fitness (CRF and S/A), cognition (UBS), and multiple path (CRF, S/A, and LBS). In boys, however, the relationship is solely mediated by the cognitive path, supporting prior research by Fernández-Sánchez et al. [11], but contrasting with findings by Lopes et al. [10], which reported no significant sex differences. The direct association between GMC and AA in girls, along with the mediation via physical fitness, might suggests a more integrated development of physical and cognitive domains in females compared to males. These differences could be based in biological and sociocultural factors that influence the development of motor skills and cognitive functions in boys and girls [38], such as the different rates of biological maturation [39] and brain development [40,41], as well as gender biases in activities performed [42]. Another possible explanation for these sex differences could be due to the fact that boys and girls use different circuits and molecular mechanisms to solve the same cognitive problems. Consequently, although general ability may be the same, it is unlikely that the strategies used by boys and girls rely on identical neurobiological mechanisms [43]. Nevertheless, given the scarcity of studies with a particular focus on sex differences, more research is needed.

With evidence that the fitness path, cognitive path, and multiple path all act as mediators on the relationship between GMC and AA, the question arises as to which of these three paths is the most influential mediator of this relationship. In the total sample, the cognitive path is the predominant mediator, followed by the fitness path and the multiple path. When analyzed by sex, for boys, only the cognitive path mediates this relationship, whereas for girls, the fitness path is the most predominant mediator, followed by the cognitive path and the multiple path. Our findings suggest the existence of a greater preponderance of the cardiovascular hypothesis [44] in the mediating mechanisms in girls, while the cognitive hypothesis [45,46] better explains these mechanisms in boys. This could be due to significant sex differences previously mentioned, especially at 9-10 years, an age at which these differences are more pronounced [47]. These different underlying mechanisms suggest that the impact of GMC on enhancing AA may vary depending on the type of physical activity, specifically whether it is cognitively engaging versus oriented towards fitness.

Finally, our results suggest that CRF and S/A act as significant mediators of cognitive benefits, while the role of muscular strength remains unclear. This could be due to different biological and neurological mechanisms described in the literature. Specifically, CRF is associated with angiogenesis in the motor cortex and increased cerebral blood flow, which enhances brain vascularization and could positively impact cognitive performance [48]. Improvements in S/A could enhance attention [49] and speed nerve impulse conduction, influencing brain processing speed [15]. These factors may improve the spinal cord function, causing synaptogenesis, increasing the BDNF and the reorganization of movement representations within the motor cortex [50]. This combined set of neural changes could positively impact on cognition [50] and, subsequently, on AA [15]. However, the relationship between muscle strength and cognition and AA remains equivocal. While some studies report positive correlations [51,52], others find no significant relationship [53,54]. Therefore, further research is needed to clarify the influence of physical fitness components on cognitive and academic outcomes.

### Limitations

The present study has some limitations to be described. First, its cross-sectional design limits our ability to infer causality. Longitudinal studies are needed to establish causal relationships and to understand the developmental trajectories of these variables. Nevertheless, the temporality proposed in our models is consistent with the current knowledge about the relationship between GMC, fitness, EF, and AA [5,9,11,55,56]. Second, the reliance on school-based assessments of AA may not capture the full spectrum of academic skills. Although mathematics and language in primary education are key cross-curricular areas for the remaining subjects and other academic competences. Third, the study does not consider potential behavior-related confounders, moderators, or mediators such as physical activity or sleep. Finally, even though the analysis in this study was controlled for some potential confounders, neither visual acuity nor adiposity was controlled for, and both are known to be health conditions affecting the performance in motor skills tests [57–59].

### Conclusions

Our results suggest that a significant proportion of the effect found between GMC and AA in children is due to improvements in fitness (CRF and S/A) and EF. These findings highlight the need for motor skills programs that target both fitness (particularly CRF and S/A) and EF to improve AA. Furthermore, the sex differences observed in the mediation pathways highlight the need for sex-sensitive approaches in interventions. Boys may benefit from engaging in activities that combine physical activity with high cognitive demands, such as perceptual-motor tasks. Conversely, girls may benefit from prioritizing activities that focus on improving physical fitness.

## Data Availability

The data underlying the results presented in the study are available upon reasonable request to the corresponding author

## Abbreviations

MC: motor competence
GMC: gross motor competence
EF: executive function
CR: cardiorespiratory fitness
S/A: speed/agility
UBS: upper body strength
LBS: lower body strength
AA: academic achievement
IE: indirect effect.

## ACKNOWLEDGMENTS

The authors thank the schools, families, and children for their enthusiastic participation in the study. We thank all membership of the Cuenca Study Group, who helped to make this study possible: Carlos Berlanga-Macías, Blanca Notario-Pacheco, María Jesús Pardo-Guijarro, Celia Álvarez-Bueno, Marta Nieto-López, Alberto González-García, Jorge Cañete García-Prieto, Ana Torres-Costoso, Antonio García-Hermoso, Caterina Pesce, and Ricardo Cuevas-Campos.

